# Cardiovascular Risk as a Moderator of the Relationship of Plasma Alzheimer’s Disease Biomarkers with Cognitive Status

**DOI:** 10.1101/2025.06.27.25330453

**Authors:** Angelina Kancheva, Donald Lyall, Kamen A. Tsvetanov, Ivana Kancheva, Kalliopi Mavromati, Ivan Koychev, Benjamin Tari, Daniela Jaime Garcia, Lynne Hughes, Joanna Wardlaw, Terence Quinn

## Abstract

**Objectives:** Plasma biomarkers may assist with diagnosis and prognosis of Alzheimer’s disease and other dementias. Cardiovascular risk is associated with impaired cognitive health, although mechanisms are not completely understood. We sought to explore whether cardiovascular risk moderates the relationship between plasma Alzheimer’s disease biomarkers and cognitive status.

**Methods:** We included groups of cognitively normal (n=301) and combined mild cognitive impairment or probable Alzheimer’s disease (n=444), based on clinical assessment, from the Bio-Hermes-001 study. Cardiovascular risk was quantified using the Atherosclerotic CVD (ASCVD) risk calculator. We conducted a series of logistic regression analyses to evaluate the association of cardiovascular risk, each of several Alzheimer’s disease biomarkers (i.e., plasma amyloid beta (Aβ)42/Aβ40, phosphorylated tau (p-tau)181, p-tau217, and circulating levels of apolipoprotein E (ApoE4)), with cognitive status. We tested moderation by cardiovascular risk in each model.

**Results:** We included 745 participants (mean age=72.3 years; 423 (56.8%) female) in the analysis. In each model, plasma biomarkers and cardiovascular risk were independently associated with cognitive status; the strongest association was found with p-tau217 (odds ratio (OR)=2.33; 95% confidence intervals (95%CI) [1.89-2.9]; *p*<0.0001). CVD risk only marginally moderated the relationships between p-tau181 and cognitive status, and between p-tau217 and cognitive status (*p*<0.05).

**Discussion:** Plasma Alzheimer’s disease biomarkers and cardiovascular risk were independently associated with cognitive status, but cardiovascular risk only marginally moderated the p-tau181- and p-tau217-cognitive status relationships. If plasma biomarkers and CVD risk potentially confer an independent risk of dementia, cardiovascular risk assessment should complement other dementia biomarker assessments in clinical and research cognitive screening.

## Introduction

Accurate detection and prognosis is crucial for individuals with cognitive complaints, and precision phenotyping of underlying neuropathology is necessary to enable assessment of disease-modifying therapies^1^. The use of biomarkers has become increasingly important in the dementia assessment process, as they constitute an objective measure of pathophysiology *in vivo*^2^. Cerebrospinal fluid (CSF)- and positron emission tomography (PET)-based measures can offer insights into neuropathology, however, the application of these techniques remain limited due to the perceived invasiveness of lumbar punctures, and low availability and high cost of PET imaging, respectively^3^. This has led to a growing interest in blood plasma biomarkers, which might be a more practical and cost-effective alternative to improve early diagnosis and help reliably predict disease risk^4^.

Plasma biomarkers may be useful in detecting varying levels of Alzheimer’s disease (AD) pathology corresponding to different stages of the clinical continuum of disease^1^. Combinations of pathology-specific biomarkers (e.g., in preclinical AD, plasma p-tau217 or the plasma Aβ42/Aβ40 ratio) with generic markers of neurodegeneration and/or neuroinflammation, such as glial fibrillary acidic protein (GFAP)^5^, demonstrate potential to identify and categorize pathological states. P-tau217 is specific to AD pathology, accurately predicts brain Aβ and tau concentrations, and reflects the topographic magnitude of tau aggregation^6^. However, no combination of biomarkers can perfectly discriminate clinical cognitive status, and other, non-amyloid and non-tau, factors likely play an important part in disease progression and presentation.

Cardiovascular disease (CVD) risk factors, including hypertension, smoking, and presence of atrial fibrillation, have a well-established association with the common cognitive syndromes seen in older adults including AD^7,8^. Cerebrovascular lesions, such as lacunes and white matter hyperintensities (WMHs), often accompany cognitive decline and AD^9,10,11^. This could be due to shared risk factors between CVD and AD and/or a potential direct pathogenic pathway from vascular pathology to neuronal loss and dementia^10^.

Despite recent advances in the biomarker field, there has been limited focus on disentangling the relationship between CVD risk and AD biomarkers. Given the high prevalence of comorbid CVD in people living with AD and ongoing development of ultra-sensitive assays for the detection of AD pathology in blood, exploring the CVD-AD biomarker relationship could improve our understanding and application of screening.

In this study, we aimed to 1) evaluate the association of select plasma AD biomarkers (plasma Aβ42/Aβ40, p-tau181, p-tau217, and circulating ApoE4) and CVD risk with cognitive status; and 2) test moderation of the relationship between each biomarker and cognitive status by CVD risk.

## Methods

### Bio-Hermes-001 Study

The purpose of the Bio-Hermes-001 study (BH001)^12^ was to interrogate the relationship of novel blood and digital biomarkers with PET-derived brain amyloid status^12^. Data for this study were provided by the study sponsor Global Alzheimer Platform (GAP) Foundation and accessed as part of the Scottish Data Challenge using an Alzheimer’s Disease Data Initiative (ADDI) workspace (https://community.addi.ad-datainitiative.org/). The Bio-Hermes cohort consists of 1,001 participants recruited from community-based populations between April 2021 and November 2022 across 17 research sites in the United States of America^12^. To ensure sufficient inclusion of underrepresented communities and individuals presenting as cognitively normal (CN), mild cognitive impairment (MCI), or mild AD, stratification was closely monitored by GAP.

### Participants and Eligibility Criteria

We included CN individuals and individuals with MCI or mild AD (combined) from the BH001 study^12^. To be included in the original BH001 study^12^, participants had to 1) be between 60 and 85 years of age (inclusive), 2) be fluent in the language that the allocated study site used (i.e., English or Spanish), 3) have a Mini-Mental State Examination (MMSE) score^13^ of 20 to 30 (inclusive) except for some instances where the principal investigator (PI) was allowed to include MMSE scores of 17 if the patient was determined to have mild AD based on clinical judgement, 4) have a normal Geriatric Depression Scale (GDS) score^14^, 5) have no known negative brain amyloid PET scan within the past 12 months, 6) have no history of stroke or seizures within 12 months, and 7) have no history of cancer within the past 5 years, excluding melanoma skin or prostate cancer *in situ*. Exclusion criteria included medical conditions that may affect cognitive assessments or preclude participation in a therapeutic trial. We did not further exclude any participants. All participants provided informed consent prior to any study procedures. The study protocol was reviewed and approved by Advarra, a central institutional review board (Reference Number Pro00046018)^12^.

### Study Groups

Participants who met eligibility criteria were allocated to three groups based on clinical screening procedures. The groups represented the range of cognitive function typically observed in clinical trials, namely CN, MCI, or mild AD.

### Cognitively Normal

The CN group consisted of participants who had no self-or partner-reported memory loss or concerns; an MMSE score^13^ of 26 to 30 (inclusive); Rey Auditory Verbal Learning Test (RAVLT) delayed recall score^15^ within a normal range based on age and sex, and no evidence of functional impairment based on the Functional Activities Questionnaire (FAQ) score^16^ in the investigator’s judgement.

### Mild Cognitive Impairment

The MCI group consisted of participants who were diagnosed with MCI based on the National Institute on Aging (NIA)-Alzheimer’s Association (AA) criteria^17^, and this was verified via medical records, or an MMSE score^13^ of 24 to 30 (inclusive); RAVLT delayed recall score^15^ of at least 1 standard deviation (SD) below the age- and race-adjusted mean; and minimal-to-mild functional impairment with preservation of independence in functional abilities in the investigator’s judgement.

### Mild Alzheimer’s Disease

The mild AD group consisted of participants who had been diagnosed with probable AD based on the NIA-AA criteria^17^ and verified through medical records), or an MMSE score^13^ of 20 to 24 (except for a few instances where the PI was allowed to include MMSE scores of 17 based on clinical judgement); RAVLT delayed recall score^15^ ≥ 1 SD below the age- and race-adjusted mean; and evidence of functional decline and loss of independence in functional abilities in the investigator’s judgement.

To increase statistical power and model stability, we combined participants with MCI (n=235) and probable AD (n=209) into one group, hereafter referred to as the cognitive impairment group (total n=444). This allowed us to focus on the presence versus absence of cognitive impairment, simplifying interpretation while retaining meaningful group differences.

### Study Visits

Participants completed three study visits conducted by the BH001 study sites^12^. During the first visit, information on participant demographics, vital signs, medical history, and concomitant medication was collected, and cognitive testing was performed. At visit two, participants underwent a brain amyloid PET scan or a lumbar puncture for CSF collection if the local site did not have access to a qualified PET imaging center. During the third visit, blood samples were collected, and brain amyloid/CSF results were reviewed.

### Blood Biospecimen Collection and Data Processing

Biomarkers of interest for the BH001 study included Aβ40, Aβ42, Aβ42/Aβ40, p-tau181, p-tau217, total (t-) tau, ApoE ε4, neurofilament light (NfL), and GFAP^12^. Plasma samples were sent to C_2_N Diagnostics laboratories for Aβ40, Aβ42, Aβ42/Aβ40, and ApoE ε4 (PrecivityAD) assays^18,19^; Quanterix laboratories for analysis of Aβ40, Aβ42, Aβ42/Aβ40, NfL, GFAP, t-tau, and p-tau181^20^; Eli Lilly and Company Clinical Diagnostics Laboratory for measurement of p-tau217^21^; and appropriate laboratories for genome and ribonucleic acid (RNA) sequencing and proteomics^12^.

To minimize potential confounding effects arising from study site variability in sample handling and measurement protocols, we focused our analysis on biomarker data from Roche Diagnostics (Aβ42, Aβ40, p-tau181, and circulating ApoE4) and Eli Lilly (p-tau217) only. Full details of the biospecimen collection protocol are available elsewhere^12^.

### Data Preprocessing

Data preprocessing steps included removing missing data, imputing implausible values as appropriate, creating an Aβ42/Aβ40 ratio variable from the Roche Diagnostics dataset (in pg/mL units), log-or square-root transforming variables if skewed, standardizing and scaling continuous variables before running any logistic regression analyses, and centering predictor variables before creating interaction terms. Full details are available in Supplementary Methods, eAppendix 1.

### Cardiovascular Disease Risk Score Calculation

We used the American College of Cardiology (ACC) and American Heart Association (AHA) Atherosclerotic Cardiovascular Disease (ASCVD) risk calculator to predict risk of future CVD^22^. The risk score considers age, sex (male or female), ethnicity (White, Black, Asian, or Other), total and high-density lipoprotein (HDL) cholesterol (in mmol/L), systolic blood pressure (SBP), information on hypertension treatment (yes or no), all-cause diabetes status (yes or no), and current smoking status (yes or no). The ASCVD risk score is suitable for predicting a first CVD event, i.e., myocardial infarction, stroke, or death due to coronary heart disease or stroke.

ASCVD risk scores were calculated using the RiskScorescvd R package (https://cran.r-project.org/web/packages/RiskScorescvd/index.html). Data transformations required for the calculation were as follows: convert HDL and total cholesterol units from mg/dL to mmol/L; transform the ethnicity variable such that it only contains four categories (i.e., White, Black, Asian, or Other); and create a sex variable based on reported sex. We conducted all analyses using continuous ASCVD scores. We calculated categorical ASCVD scores for descriptive purposes as follows: very low risk of atherosclerosis (<5%), low risk (5% to < 10%), moderate risk (10% to < 20%), or high risk (≥20%), as per RiskScorescvd R package.

### Statistical Analysis

#### Regression Analyses

All analyses were conducted using the R statistical software (version 4.4.1). To explore the relationship between plasma AD biomarkers, ASCVD, and cognitive impairment, we first ran four logistic regression analyses. Model formulas using Wilkinson’s notation were specified as follows: Model 1:*‘Cognitive status* ∼ *Aβ42/Aβ40* + *ASCVD score’*; Model 2:*‘Cognitive status* ∼ *p-tau181* + *ASCVD score’*; Model 3:*‘Cognitive status* ∼ *p-tau217* + *ASCVD score’*; Model 4:*‘Cognitive status* ∼ *circulating ApoE4* + *ASCVD score’*. We then tested moderation by CVD risk in each model; Model 1*i*:*‘Cognitive status* ∼ *Aβ42/Aβ40*\**ASCVD score’*; Model 2*i*:*‘Cognitive status* ∼ *p-tau181*\**ASCVD score’*; Model 3*i*:*‘Cognitive status* ∼ *p-tau217*\**ASCVD score’*; Model 4*i*:*‘Cognitive status* ∼ *circulating ApoE4*\**ASCVD score’*.

We calculated Akaike Information Criterion (AIC), pseudo R², and Area Under the Curve (AUC) values to assess model fit for each model. We report odds ratios (OR), 95% confidence intervals (95%CI), and unadjusted *p*-values denoting statistical significance for each model. We chose to present unadjusted estimates since we were primarily interested in the raw associations and a predefined objective, i.e., to test moderation of the relationship between plasma AD biomarkers and cognitive status by CVD risk. Hereafter, we refer to associations surviving a *p*<.001 (unadjusted) threshold as statistically significant and associations surviving a *p*<.05 threshold (unadjusted) as marginally significant.

#### Additional Analyses

To explore potential differences between components of the ASCVD risk score, we ran separate logistic regression analyses with each component and each biomarker of interest as predictors and cognitive impairment as the outcome variable. Additionally, where the interaction term between biomarker of interest and ASCVD risk score was at least marginally significant (i.e., at *p*<0.05), we split the data according to sex and re-ran the analyses on males and females separately.

We further split the data into an incident CVD group (i.e., ongoing or history of major adverse CVD events, stroke, angina, peripheral vascular disease, or heart disease; all derived from participants’ medical history) and a no CVD group and repeated all analyses. We conducted these analyses for exploratory purposes.

## Results

### Participant Characteristics

Participants’ mean (standard deviation; SD) age was 72.3 (6.6) and 423 (56.8%) were female. Mean (SD) SBP was 133.2mmHg (16.06), mean (SD) diastolic BP was 77.3mmHg (9.01), and 58 (7.8%) of the sample had diabetes. In terms of CVD risk, the majority of participants (n=372; 49.9%) had a high risk of ASCVD (see Supplementary Material, eAppendix 2). Baseline participant characteristics are reported in Table 1.

**Table 1.** Description of the Bio-Hermes cohort included in the analysis (n=745).

| **Bio-Hermes Variable** | **Mean (SD)** |
| --- | --- |
| **Age (years)** | 72.3 (6.6) |
| **Sex, number (%)** |  |
| Female | 423 (56.8) |
| Male | 322 (43.2) |
| **Systolic Blood Pressure (mmHg)** | 133.2 (16.1) |
| **Diastolic Blood Pressure (mmHg)** | 77.3 (9.0) |
| **Diabetes status, number (%)** |  |
| Yes | 58 (7.8) |
| No | 687 (92.2) |
| **Current smoking, number (%)** |  |
| Yes | 40 (5.4) |
| No | 705 (94.6) |
| **Ethnicity, number (%)** |  |
| Not Hispanic or Latino | 655 (87.9) |
| Hispanic or Latino | 80 (10.7) |
| Not Reported | 10 (1.3) |
| **Ongoing medication for hypertension, number (%)** |  |
| Yes | 410 (55.0) |
| No | 335 (45.0) |
| **Baseline ASCVD Risk Classification, number (%)** |  |
| Very low risk | 49 (6.6) |
| Low risk | 78 (10.5) |
| Moderate risk | 246 (33.0) |
| High risk | 372 (49.9) |

*Abbreviations:* ASCVD, atherosclerotic cardiovascular disease; SD, standard deviation.

### Association of plasma AD Biomarkers and CVD Risk with Cognitive Impairment

Lower Aβ42/Aβ40 values (OR=0.84; 95%CI [0.72-0.98]) and higher ASCVD risk (OR=1.63; 95%CI [1.39-1.91]) were associated with an increased probability of cognitive impairment in Model 1 (AIC=962.13; pseudo R²=0.05).

Higher p-tau181 levels (OR=1.83; 95%CI [1.53-2.18]) and higher ASCVD risk (OR=1.43; 95%CI [1.22-1.67]) were associated with an increased probability of cognitive impairment in Model 2 (AIC=917.31; pseudo R²=0.09).

Higher p-tau217 levels (OR=2.33; 95%CI [1.89-2.90]) and higher ASCVD risk (OR=1.47; 95%CI [1.24-1.76]) were associated with an increased probability of cognitive impairment in Model 3 (AIC=883.55; pseudo R²=0.13).

Finally, higher circulating levels of ApoE4 (OR=1.33; 95%CI [1.13-1.55]) and higher ASCVD risk (OR=1.70; 95%CI [1.45-1.99]) were associated with an increased probability of cognitive impairment in Model 4 (AIC=954.61; pseudo R²=0.06). Table 2 details the results from the logistic regression main effects models.

**Table 2.** Logistic regression main effects models^a^.

| Variable | P-value | OR | 95%CI | Pseudo R<sup>2</sup> | AIC | AUC |
| --- | --- | --- | --- | --- | --- | --- |
| **<u>Model 1: Aβ42/Aβ40 + ASCVD risk</u>** |  |  |  |  |  |  |
|  |  |  |  | 0.05 | 962.13 | 0.65 |
| Aβ42/Aβ40 | 0.02 | 0.84 | 0.72-0.98 | | | |
| ASCVD Risk Score | <0.0001 | 1.63 | 1.39-1.91 |  |  |  |
| **<u>Model 2: P-tau181 + ASCVD risk</u>** |  |  |  |  |  |  |
|  |  |  |  | 0.09 | 917.31 | 0.71 |
| P-tau181 | <0.0001 | 1.83 | 1.53-2.18 |  |  |  |
| ASCVD Risk Score | <0.0001 | 1.43 | 1.22-1.67 |  |  |  |
| **<u>Model 3: P-tau217 + ASCVD risk</u>** |  |  |  |  |  |  |
|  |  |  |  | 0.13 | 883.55 | 0.73 |
| P-tau217 | <0.0001 | 2.33 | 1.89-2.90 |  |  |  |
| ASCVD Risk Score | <0.0001 | 1.47 | 1.24-1.76 |  |  |  |
| **<u>Model 4: ApoE4 + ASCVD risk</u>** |  |  |  |  |  |  |
|  |  |  |  | 0.06 | 954.61 | 0.66 |
| ApoE4 | <0.001 | 1.33 | 1.13-1.55 |  |  |  |
| ASCVD Risk Score | <0.0001 | 1.71 | 1.45-1.99 |  |  |  |

*Abbreviations:* AIC, Akaike Information Criterion; ASCVD, atherosclerotic cardiovascular disease; AUC, area under the curve; CI, confidence interval; OR, odds ratio; SE, standard error.
<sup>a</sup>In each model, the respective biomarker and ASCVD risk score were introduced as independent variables and CI as dependent variable.

### Moderation by CVD Risk

ASCVD risk did not significantly moderate the relationship between Aβ42/Aβ40 and cognitive impairment (Model 1*i* AIC=962; pseudo R²=0.05; *p*=0.14), and ApoE4 and cognitive impairment (Model 4*i* AIC=956.36; pseudo R²=0.06; *p*=0.61). ASCVD risk only marginally moderated the relationships between p-tau181 and cognitive impairment (Model 2*i* AIC=914.15; OR=0.81; 95%CI [0.68-0.97]; pseudo R²=0.10), and p-tau217 and cognitive impairment (Model 3*i* AIC=880.39; OR=0.78; 95%CI [0.63-0.97]; pseudo R²=0.13), both *p*=0.02. Table 3 details the results from the logistic regression interaction effects models.

**Table 3.** Logistic regression interaction effects models^a^.

| Variable | P-value | OR | 95%CI | Pseudo R<sup>2</sup> | AIC | AUC |
| --- | --- | --- | --- | --- | --- | --- |
| **<u>Model 1i:</u> Aβ42/Aβ40 + ASCVD risk + moderation by ASCVD risk** |  |  |  |  |  |  |
|  |  |  |  | 0.05 | 962 | 0.65 |
| **Aβ42/Aβ40** | 0.03 | 0.84 | 0.72-0.98 |  |  |  |
| **ASCVD Risk Score** | <0.0001 | 1.62 | 1.38-1.90 |  |  |  |
| **Aβ42/Aβ40 x ASCVD Risk Score** | 0.14 | 1.13 | 0.97-1.32 |  |  |  |
| **<u>Model 2i:</u> P-tau181 + ASCVD risk + moderation by ASCVD risk** |  |  |  |  |  |  |
|  |  |  |  | 0.10 | 914.15 | 0.72 |
| **P-tau181** | <0.0001 | 1.79 | 1.50-2.13 |  |  |  |
| **ASCVD Risk Score** | <0.0001 | 1.40 | 1.18-1.68 |  |  |  |
| **P-tau181 x ASCVD Risk Score** | 0.02 | 0.81 | 0.68-0.97 |  |  |  |
| **<u>Model 3i:</u> P-tau217 + ASCVD risk + moderation by ASCVD risk** |  |  |  |  |  |  |
|  |  |  |  | 0.13 | 880.39 | 0.74 |
| **P-tau217** | <0.0001 | 2.36 | 1.90-2.94 |  |  |  |
| **ASCVD Risk Score** | <0.001 | 1.38 | 1.16-1.64 |  |  |  |
| **P-tau217 x ASCVD Risk Score** | 0.02 | 0.78 | 0.63-0.97 |  |  |  |
| **<u>Model 4i:</u> ApoE4 + ASCVD risk + moderation by ASCVD risk** |  |  |  |  |  |  |
|  |  |  |  | 0.06 | 956.36 | 0.66 |
| **ApoE4** | <0.001 | 1.33 | 1.14-1.56 |  |  |  |
| **ASCVD Risk Score** | <0.0001 | 1.71 | 1.47-2.01 |  |  |  |
| **ApoE4 x ASCVD Risk Score** | 0.61 | 1.05 | 0.88-1.25 |  |  |  |

*Abbreviations:* AIC, Akaike Information Criterion; ASCVD, atherosclerotic cardiovascular disease; AUC, area under the curve; CI, confidence interval; OR, odds ratio; SE, standard error.
<sup>a</sup>In each model, the interaction term between the respective biomarker and ASCVD risk score was introduced to assess moderation by CVD risk.

### Additional Analyses

Across models, increasing age and male sex (versus female) were the two factors consistently associated with an increased risk of cognitive impairment. For all associations, see Supplementary Material, eAppendices 3-6.

Additionally, in the analyses split according to sex, higher p-tau181 was associated with an increased probability of cognitive impairment for both males and females (for males, OR=1.67; 95%CI [1.27-2.19]; for females, OR=1.97; 95%CI [1.56-2.50]). However, higher ASCVD risk only related to a higher probability of cognitive impairment for females (OR=1.51; 95%CI [1.21-1.87]), but not males. In interaction effects models, ASCVD risk only marginally negatively moderated the relationship between p-tau181 and cognitive impairment for males (OR=0.68; 95%CI [0.49-0.94]), but not females. A similar pattern of results was observed for p-tau217 (for full details, see Supplementary Material, eAppendices 7-14).

In all CVD-stratified analyses, only higher ASCVD risk was associated with cognitive impairment for participants with incident CVD. For participants with no CVD, both ASCVD risk score and biomarker of interest were associated with cognitive impairment. For full details, see Supplementary Material, eAppendices 15-26.

## Discussion

Using data from the BH001 study, we evaluated the cross-sectional association between several plasma AD biomarkers (plasma Aβ42/Aβ40, p-tau181, p-tau217, and circulating ApoE4) and CVD risk, with cognitive status, and tested moderation of the relationship between each biomarker and cognitive status by CVD risk. Plasma AD biomarkers and CVD risk were significantly independently associated with cognitive status. Across biomarkers, the strongest association was with p-tau217. CVD only marginally moderated the relationship of p-tau181 and p-tau217 with cognitive status.

### Plasma AD Biomarkers and Cognitive Status

In the original BH001 study^12^, plasma Aβ42/Aβ40, p-tau181, and p-tau217 predicted brain amyloid positivity, and p-tau217 reported superior discrimination of cognitive impairment (i.e., the p-tau217 AUC was larger than Aβ42/Aβ40 and p-tau181). We extend these findings by confirming the importance of both AD biomarkers and CVD risk for cognitive impairment. Plasma AD biomarkers have consistently shown excellent correspondence with CSF or PET biomarkers^12,23^ but their clinical utility remains an active area of research. In our analysis, the Aβ42/Aβ40 ratio had the weakest association with concurrent cognitive impairment. In a recent study using data from the MEMENTO cohort^23^, plasma Aβ42/Aβ40 was less efficient than CSF Aβ42/Aβ40 in predicting AD conversion risk, whereas blood and CSF p-tau181 had similar risk prediction abilities. Notably, peripheral tissues might contribute to both circulating amyloid and AD pathology in the brain and cerebral vasculature, suggesting that amyloid sources outside the central nervous system (CNS) should be accounted for when assessing Aβ levels in blood^24^. Taken together, these findings indicate that more research is needed to assess the diagnostic utility of plasma Aβ42/Aβ40.

Regarding plasma tau biomarkers, p-tau217 was positively associated with cognitive impairment most strongly in our analysis, in line with previous work^12,25^. P-tau217 has been shown to have higher discriminative accuracy than p-tau181 and magnetic resonance imaging (MRI) measures to differentiate clinically diagnosed AD from other neurodegenerative conditions but is not superior to CSF p-tau181 and CSF p-tau217^21^. One explanation for this could be that plasma p-tau217 levels start to decrease earlier in the disease process compared to plasma p-tau181^25^, but more research is needed to confirm this.

Studies on the efficacy of plasma tau in AD diagnosis have thus far provided contradictory results with evidence for elevated, mild, legible, and reduced levels in AD^26,27,28,29^, respectively. In memory clinics, plasma p-tau181, plasma p-tau217, and plasma p-tau231 also differentially map onto brain amyloid load and tau aggregation^30^. Across studies, plasma tau appears to reliably and accurately reflect AD pathology^40^. However, the overlap of this signal with that seen during healthy ageing is still considerable^31,32^, and several tau markers have been shown to reflect different phases of AD progression^30^, underscoring the need for further studies on their diagnostic potential.

### CVD Risk and Cognitive Status

The link between CVD risk and cognitive impairment is well-established^33^. In our study, higher CVD risk as measured with the ASCVD risk score was independently associated with cognitive impairment in each biomarker model, in line with previous cross-sectional and longitudinal studies^7,8,33^. The strongest association was observed in the ApoE4 main effects model (Model 4), complementing previous reports that circulating ApoE4 protein might magnify lifestyle risks for dementia^34^.

Regarding moderation, CVD risk only marginally moderated the relationships between p-tau181 and p-tau217 and cognitive impairment, respectively, indicating that CVD risk and plasma biomarkers might exert independent effects on cognitive impairment. Both CVD and dementia are progressive diseases with asymptomatic pre-clinical phases, which can last decades^35^. Although increasing age is the greatest risk factor for both CVD and dementia, both conditions share multiple other modifiable risk factors including diabetes, hypertension, and smoking status^36^. Although it is not precisely established whether brain amyloid accumulation is secondary to vascular burden, there is evidence that atherosclerosis may enhance amyloid deposition through increased inflammation, hypoxia, and oxidative stress^37^. Vascular risk factors could thus predispose individuals to AD by promoting amyloid accumulation in the brain, and may manifest clinically several years earlier than AD^38^.

Research exploring the association between CVD, plasma AD biomarkers, and cognitive impairment is still scarce. There is mixed evidence as to whether CVD risk confers independent or joint risk of dementia in combination with other relevant factors^39^. Here, we provide evidence that CVD risk may contribute to cognitive impairment beyond the effects of plasma AD biomarkers. Regarding individual CVD risk factors, age and sex were consistently associated with higher risk of cognitive impairment across models, as in previous studies^7,10^. Systolic blood pressure emerged as an important factor in the p-tau217 model but the association was weak. Of note, CVD risk was calculated based on participants’ medical history, whereas biomarkers of interest reflected biological measurements and thus represent a different class of biomarker. This should be considered when interpreting our findings.

Given the extensive social and economic burden of AD globally, our findings suggest that CVD risk assessment could complement other dementia biomarker assessments. This might be particularly important for individuals who already experience a substantial CVD burden. Indeed, results from our CVD-stratified analyses revealed that, for this subgroup, only CVD risk was significantly associated with cognitive impairment. The observation that the utility of plasma biomarkers was reduced in the incident CVD subgroup emphasizes the importance of comprehensive CVD assessment and cardiovascular risk management (CVRM) alongside biomarkers of neurodegeneration and neuroinflammation. Our findings extend a recent investigation of over 10,000 individuals from the Whitehall II study^40^ where dementia risk in CVD patients was lower when lifestyle factors, such as diabetes and hypertension, were at levels complying with recommended guidelines^41^. Importantly, the benefits of CVRM may differ between persons with and without dementia^42^. This should be considered by healthcare providers when initiating or intensifying CVD treatment. Population screening for CVD risk is established in many healthcare systems, and recommended in guidelines^43^. Our data suggests the potential efficacy of combining CVD risk screening data with risk screening for cognitive impairment. The economic and opportunity cost would be minimal for those healthcare systems where CVD risk screening is already in place, and the improvements in diagnostic and prognostic utility for dementia could be substantial. Finally, our results support the incorporation of vascular burden (V) into the well-established AD amyloid-beta accumulation (A), tau aggregation (T), and neurodegeneration (N) ATN classification framework^44^.

### Strengths and Limitations

The main strength of our study is the focus on the relationship between CVD risk and novel plasma AD biomarkers in the context of cognitive impairment, an under-investigated topic. By demonstrating that the effects of plasma AD biomarkers and CVD risk on cognitive impairment might be additive rather than synergistic, we emphasize the importance of targeting modifiable CVD risk factors as a preventative dementia strategy, which constitutes an additional strength.

A noteworthy limitation is that our results cannot be used to infer causality regarding the directionality of the reported associations due to the cross-sectional design of the BH001 study. Further, the BH001 study only included participants aged 60 to 85 years of age, suggesting that our observations do not generalise to individuals outside this age range. Finally, it is likely that our sample was insufficiently powered to reveal meaningful sex differences or differences between individual ASCVD components as related to plasma AD biomarkers and cognitive impairment. Efforts to further explore these relationships should be pursued going forward.

### Implications and Future Directions

Our findings complement a growing body of research supporting the potential of plasma AD biomarkers to enhance AD detection and predict risk of dementia. Given the high cost, perceived invasiveness, and resource-intensive nature of CSF- and PET-based biomarkers, plasma biomarkers are increasingly likely to be incorporated into intervention trials and clinical practice^45^. Existing biomarkers are imperfect, and our data suggest the utility of combining neurodegenerative biomarkers with other risk assessments. Future research may wish to look for additive or synergistic effects of AD biomarkers and other potentially relevant pathologies, such as inflammation.

Regarding CVD risk, the evidence we provide of a possibly independent contribution of CVD risk to cognitive impairment underscores the importance of CVD risk monitoring. This is particularly important in the early, asymptomatic stages of AD when intervention strategies might be more effective^46^. We encourage the promotion of healthy lifestyles to increase cardiovascular and brain health by focusing on modifiable CVD risk factors and potentially help delay onset of clinically observable dementia symptoms.

Although our cross-sectional analysis precludes ascertaining cut-off points for CVD burden as related to progression from MCI to probable AD, future work should aim to elucidate the contribution of both individual CVD risk factors and cumulative vascular burden to transitioning from pre-clinical to clinical AD stages. This includes assessing the utility of various CVD risk score calculators and comparing their correspondence to assess vascular burden as related to plasma AD biomarkers. Finally, larger, longitudinal studies in diverse populations with different medical comorbidities are required to determine which plasma biomarkers are optimal in various clinical scenarios, as well as in relation to varying loads of CVD burden. Shedding light on the mechanisms linking vascular disease and AD pathology in blood should be a research priority in the future.

### Conclusions

In summary, our findings suggest that plasma biomarkers and CVD risk might confer independent risk of dementia. Given the multiple risk factors for dementia and large implications of AD prevention, treatment and management for healthcare systems worldwide, CVD risk assessments should complement other dementia biomarker assessments. Future studies using large and diverse populations are required to clarify the relationship between plasma biomarkers and CVD risk at different stages of the AD clinical continuum.

## Data Availability

All data were provided by Global Alzheimer's Platform Foundation.

## Acknowledgements

All authors acknowledge the help and support of Global Alzheimer’s Platform Foundation and Alzheimer’s Disease Data Initiative in enabling access to the Bio-Hermes-001 study data through the AD workbench. We would also like to thank Lynne Hughes, Chief of Staff at GAP Foundation, as the representative of GAP Foundation.

## Author Contributions

A.K., D.L. and T.Q. contributed to the conception and design of the study. A.K., K.M. and L.H. contributed to the acquisition of data. A.K., K.A.T., I.K., and K.M. contributed to the planning and execution of the data analysis. All co-authors contributed to the drafting and editing of the manuscript or figures.

## Sources of Funding

A.K. is supported by the Medical Research Council Doctoral Training Program in Precision Medicine (project code 314327-03).

D.L. has no funding sources relevant to the present manuscript.

K.A.T. was supported by a Fellowship Award from the Alzheimer’s Society, UK (Grant Nr. 602).

I.K.* has no funding sources relevant to the present manuscript.

K.M. has no funding sources relevant to the present manuscript.

I.K.** has no funding sources relevant to the present manuscript.

B.T. has no funding sources relevant to the present manuscript.

D.J.G. is funded by the Wellcome Trust (Grant number 224912/Z/21/Z; Translational Neuroscience Ph.D. programme at the University of Edinburgh).

Lynne Hughes is Chief of Staff at GAP Foundation.

J.W. has no funding sources relevant to the present manuscript.

T.Q. has no funding sources relevant to the present manuscript.

## Disclosures

A.K. has nothing to disclose.

D.L. has nothing to disclose.

K.A.T. has nothing to disclose.

I.K.* has nothing to disclose.

K.M. has nothing to disclose.

I.K.** has nothing to disclose.

B.T. has nothing to disclose.

D.J.G. has nothing to disclose.

Lynne Hughes is Chief of Staff at GAP Foundation.

J.W. has nothing to disclose.

T.Q. has nothing to disclose.

*I.K.* - Ivana Kancheva MSc*

*I.K.** - Dr Ivan Koychev*

## Notes

### Competing Interest Statement

The authors have declared no competing interest.

### Author Declarations

All participants provided informed consent prior to any study procedures. The study protocol was reviewed and approved by Advarra, a central institutional review board (Reference Number Pro00046018).

